# Hybrid risk scores integrating polygenic and clinical variables for endometriosis prediction

**DOI:** 10.64898/2026.08.31.26361798

**Authors:** Oksana Goroshchuk, Dora Koller

## Abstract

**Background:** Endometriosis affects approximately 10% of reproductive-age women and is associated with substantial diagnostic delay and heterogeneous symptom presentation. Prior machine-learning prediction models have relied on comorbidity data alone or on small candidate-variant genetic scores, with inconsistent or incompletely reported performance. No study has combined a well-powered, multi-ancestry polygenic risk score (PRS) with environmental, reproductive, and symptom data in a single hybrid model. We developed and evaluated hybrid risk-prediction models integrating a genome-wide, multi-ancestry PRS with clinical and symptom data for endometriosis in the US-based *All of Us* Research Program.

**Methods:** Among 69,376 participants (15,382 endometriosis cases, 53,994 controls) across six genetically inferred ancestry groups, we computed individual-level PRS values using PRS-CS weights derived from an independent, multi-ancestry GWAS. Five nested logistic regression, random forest, and XGBoost models progressively added age, ancestry, and within-ancestry genetic principal components (Model 1), environmental and reproductive factors (Model 2), symptom and comorbidity indicators (Model 3), all covariates combined (Model 4), and PRS × environment interactions (Model 5). Performance was assessed by AUROC in a held-out test set and 5-fold cross-validation, with class-weighted, Youden-optimized thresholds used for sensitivity, specificity, and predictive values; permutation importance identified top contributors. Pairwise AUROC differences were tested with a Holm-corrected DeLong-type test.

**Results:** Discrimination improved from AUROC 0.63 (PRS, age, ancestry, principal components) to 0.72 for the full model, driven mainly by symptom and comorbidity data. XGBoost consistently outperformed logistic regression and random forest. The PRS ranked among the top individual predictors by permutation importance in nearly every model, alongside age, while genetic and demographic information alone gave only modest discrimination, and PRS × environment interactions did not improve on environmental factors alone. Threshold optimization yielded balanced sensitivity and specificity (~0.67/0.65) versus near-zero sensitivity at a default threshold.

**Conclusions:** Combining the PRS with symptom and comorbidity data gave the best discrimination compared to solely a well-powered, multi-ancestry PRS as a predictor of endometriosis. This study clarifies both the promise and current limits of hybrid genetic-clinical prediction for endometriosis and points to symptom-based phenotyping, molecular subtyping, and external validation as priorities.

## Introduction

Endometriosis is a chronic systemic inflammatory disease^1^ in which endometrial-like tissue grows outside the uterine cavity^2^. The disease affects approximately 10% of reproductive-age women and girls^2^. Although its most recognized manifestations are chronic pelvic pain, dysmenorrhea, and infertility, endometriosis is increasingly understood as a systemic condition with wide-ranging effects on physical and mental health, including gastrointestinal symptoms, migraine, fatigue, anxiety, and depression^2–4^. Diagnosis has historically relied on surgical or imaging confirmation and is delayed by an average of 4-11 years from symptom onset^2^, contributing to a substantial cumulative burden on quality of life, education, work, and relationships^5^.

Part of the challenge in endometriosis detection is due to the limited disease awareness^6^, and the heterogeneity and non-specificity of negative menstrual symptoms^2^. Data-driven clustering has begun to map this heterogeneity. Using latent class analysis in the *All of Us* Research Program, we recently identified distinct symptom phenotypes, including high pain-gastrointestinal and predominantly psychological-neurological clusters, showing that the disease extends well beyond its classical gynecological presentation^3^. Other studies have derived comorbidity-based subgroups from primary-care and multi-center health records^7,8^. These studies characterize symptom and comorbidity structure but were not designed for individual-level risk prediction.

Currently, the definitive diagnosis is often delayed and requires invasive procedures. Therefore, there is growing interest in non-invasive tools that use available health information to identify women at elevated risk of endometriosis and prioritize them for evaluation. Machine-learning models trained on self-reported symptoms can classify endometriosis with high apparent accuracy. For example, a symptom-questionnaire model achieved an AUC of 0.94^9^, but such performance has typically been obtained in small, balanced samples recruited from disease-specific communities, evaluated only by internal cross-validation. Being trained against undiagnosed rather than clinically confirmed controls, this model left the real-world generalizability uncertain. In a large, population-based electronic health record cohort, discrimination has been more modest: a comorbidity-based model in approximately 628,000 Spanish primary-care records achieved an AUC of 0.73, but the model was highly unbalanced with a low positive predictive value of 1.5%^10^.

Endometriosis also has a substantial genetic component, with a twin-based heritability of approximately 50%^11^. Our recent genome-wide association study (GWAS) identified 80 associated loci and enabled the construction of multi-ancestry polygenic risk scores (PRS)^12^. However, PRS are only modestly predictive and are not yet suitable for clinical risk stratification, particularly across diverse populations^12^. Where genetic information has been incorporated into prediction models, it has comprised a limited set of candidate GWAS variants rather than a genome-wide PRS, and has added little. In the UK Biobank, a model combining lifestyle, physical, and ICD-10 diagnostic features reached an AUC of 0.80 (reported without specificity, sensitivity or positive and negative predictive values), and adding 65 or 399 candidate variants did not improve it, with a candidate-variant score alone failing to separate cases from controls^13^. A few studies have nonetheless begun to examine how genetic risk relates to clinical presentation. Our multi-ancestry GWAS reported interactions between the endometriosis PRS and symptoms such as abdominal pain, anxiety, migraine, and nausea^12^, and another study^14^ described interactions between genetic risk and comorbid conditions, but these analyses did not translate to an integrated predictive framework. To our knowledge, no study has combined a well-powered, multi-ancestry PRS with environmental, reproductive, and symptom information in a hybrid risk-prediction model for endometriosis in an ancestrally diverse population.

Here, we developed and compared hybrid risk-prediction models for endometriosis in the multi-ancestry *All of Us* cohort, integrating an endometriosis PRS with environmental factors, reproductive history, and symptom and comorbidity data. Using three machine-learning classifiers and a common evaluation framework, we quantified the incremental discrimination contributed by each information layer, tested whether PRS interacts with environmental factors, and identified the predictors driving model performance. Our goal was to clarify the relative contributions of genetic and non-genetic factors to endometriosis risk prediction and to assess whether combining them improves discrimination in a diverse, real-world population.

## Methods

### Study Population

Data were drawn from the *All of Us* Research Program, a longitudinal cohort of more than 633,000 US adults designed to over-represent populations historically underrepresented in biomedical research. Analyses used the Controlled Tier v8 release, which integrates electronic health record (EHR) data captured between May 6, 2018 and October 1, 2023^15,16^. Endometriosis cases were ascertained from two complementary sources following prior work^3^. Self-reported cases (n=17,332) were identified from the “Personal and Family Health History” survey item “Including yourself, who in your family has had endometriosis?” (response “Self”), and clinically confirmed cases (n=7,607; surgically or non-surgically diagnosed) were identified using SNOMED code 129103003. The two sources yielded 22,438 unique female endometriosis cases. Controls were female participants without an endometriosis diagnosis. To reduce participation/responder bias, controls were restricted to individuals who responded to the family-history survey question, ensuring that cases and controls were drawn from a comparable, survey-engaged population rather than allowing systematic differences in survey completion to confound case-control comparisons. Because the present analysis is built around PRS, the sample was further restricted to participants with genetic data. The final analytic sample comprised 15,382 cases and 53,994 controls across all ancestry groups.

The 19 symptom and comorbidity variables, socioeconomic and quality-of-life measures were defined in our prior work^3^. Briefly, symptoms and comorbidities (abdominal pain, anxiety, bloating, chronic pelvic pain, constipation, depression, diarrhea, dysmenorrhea, dyspareunia, dysuria, eating disorders, chronic fatigue, fibromyalgia, irritable bowel syndrome, infertility, low back pain, menorrhagia, migraine, and nausea) were coded as binary indicators from self-reported and EHR-derived data (Supplemental Table). Body mass index (BMI) was summarized as the median across each participant’s available measurements, with implausible values (<10 or >100 kg/m^2^) set to missing. Contraceptive use was derived from the drug exposure table and coded as a binary indicator (≥1 recorded contraceptive medication vs. none). Area-level socioeconomic status was captured with a ZIP-code-level deprivation index linked to each participant’s residential ZIP code. Family history of endometriosis was included as a three-level variable from the “Personal and Family Health History” survey item “Including yourself, who in your family has had endometriosis?”. Participants endorsing an affected relative (mother, sibling, daughter, or grandmother) were coded as 1, survey responders not endorsing any affected relative as 0, and non-responders as missing.

### Genetic risk score construction

The generation and quality control of *All of Us* whole-genome sequencing data have been described in detail elsewhere^17^. *All of Us* participants were assigned to one of six genetically inferred ancestry groups: African (AFR), Admixed American (AMR), East Asian (EAS), European (EUR), Middle Eastern (MID), and South Asian (SAS) using a random forest classifier trained on principal components of the Human Genome Diversity Project and 1000 Genomes reference panels and following the gnomAD ancestry labels, as previously described^17^. Endometriosis polygenic risk scores (PRS) were computed separately within each ancestry group, as described in our previous multi-ancestry endometriosis GWAS^12^. Briefly, posterior variant effect sizes were estimated with PRS-CS^18^ applied to the European-ancestry endometriosis GWAS meta-analysis, using the 1000 Genomes European population as the linkage-disequilibrium reference panel, and per-participant scores were computed with PLINK2^19^. To preserve independence between the discovery and target samples, the European target was scored using weights from a meta-analysis that excluded *All of Us*. For the non-European groups, the European-derived weights were applied to non-overlapping individuals. Participants without a valid PRS after quality control were excluded from PRS-based analyses. PRS were standardized within each ancestry group, then merged into a single multi-ancestry analytic dataset that retained all participants. The first ten within-ancestry principal components were included as covariates in all models to adjust for residual population stratification.

### Prediction model development

Missing values in race, BMI, family history of endometriosis, and deprivation index were imputed using multiple imputation by chained equations (MICE), with predictive mean matching for the continuous variables (BMI, deprivation index) and logistic regression for the binary variables (family history of endometriosis). Imputation models used predictors age, PRS, ancestry, family history, contraceptive use for BMI, age, PRS, ancestry, BMI, family history, contraceptive use for deprivation index, and age, PRS, ancestry, BMI, deprivation index, contraceptive use for family history. Race, as it had a missingness higher than 5% in the sample was excluded from the models based on previous recommendations and was retained only for cohort description. Five models were compared, all sharing a common base of PRS, age, genetically inferred ancestry, and the first ten principal components. Model 1 comprised this genetic and demographic base alone. Model 2 added environmental and reproductive factors (BMI, deprivation index, contraceptive use, and family history of endometriosis). Model 3 added the 19 symptom and comorbidity indicators to the base. Model 4 included all base, environmental/reproductive and symptom variables. Model 5 extended the base with environmental/reproductive factors and their interactions with PRS (PRS × BMI, PRS × deprivation index, PRS × contraceptive use, and PRS × family history). Ancestry was treated as a categorical variable and one-hot encoded before model fitting, and continuous predictors were entered without further transformation. For each model, three classifiers were compared: logistic regression, random forest, and XGBoost. Data were partitioned into training and held-out test sets using an 80:20 stratified split that preserved the case-control ratio, and models were additionally evaluated by 5-fold stratified cross-validation, with predictions generated out of fold. Because the outcome was imbalanced, both unweighted and class-weighted versions of each model were evaluated (class weights for logistic regression and random forest, positive-class weighting for XGBoost), with unweighted models designated the primary analysis and class-weighted models a sensitivity analysis. Model performance was assessed at the default probability threshold of 0.5 and at a threshold maximizing the Youden J statistic, determined separately for each model. The area under the receiver operating characteristic curve (AUROC) was the primary discrimination metric and sensitivity, specificity, positive and negative predictive value, balanced accuracy, F1 score, and confusion-matrix counts were also reported, for both the held-out test set and the cross-validated predictions. Discrimination was compared across the five models using all ten pairwise AUROC comparisons on the same individuals, based on out-of-fold cross-validated predictions and a DeLong-type test for correlated ROC curves, with Holm correction. Predictor contributions for the best-performing model were assessed by permutation importance, defined as the mean decrease in AUROC over 20 permutations of each feature in the held-out test set. Data curation, quality control, and imputation were performed in RStudio and predictive modeling and model comparison were performed in Python JupiterLab, all within the *All of Us* Researcher Workbench.

## Results

### Study Population

The analytic sample comprised 69,376 women (15,382 endometriosis cases and 53,994 controls) across six genetic ancestry groups, most of European ancestry (74.8%), followed by Admixed American (11.1%) and African (10.6%) (Table 1). Cases and controls were similar in age (57.5±14.4 vs 56.8±16.7 years) and area-level deprivation (0.31±0.06 in both groups). Although both reached statistical significance given the large sample size, the differences were negligible in magnitude. Body mass index was higher in cases than controls (30.5±7.8 vs 29.0±7.6 kg/m^2^, p=8.1×10^-93^). The standardized endometriosis PRS was substantially higher in cases than controls (0.25±1.01 vs −0.05±1.00, p=1.6×10^-237^), consistent with the score reflecting genetic liability to endometriosis in this cohort. All 19 symptom and comorbidity indicators were significantly more frequent in cases than in controls (Table 1). The largest relative differences were observed for dysmenorrhea (11.6% vs 3.0%), chronic pelvic pain (32.2% vs 17.5%), fibromyalgia (17.1% vs 7.0%), menorrhagia (10.1% vs 4.5%), and infertility (5.2% vs 2.3%). The most prevalent symptoms, such as anxiety (53.6% vs 39.7%), depression (53.9% vs 41.4%), abdominal pain (48.5% vs 32.8%), and migraine (43.3% vs 26.5%) were also more common among cases. Family history of endometriosis was reported more than twice as often by cases (21.1% vs 9.5%, p = 1.1×10^-253^), and contraceptive use was more common among cases (32.9% vs 25.5%). Ancestry and self-identified race distributions differed modestly between groups (for example, East Asian ancestry was less frequent among cases, 1.4% vs 2.5%).

**Table 1.**
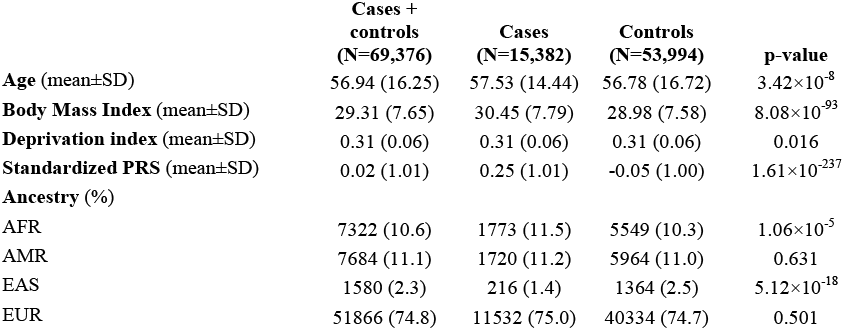
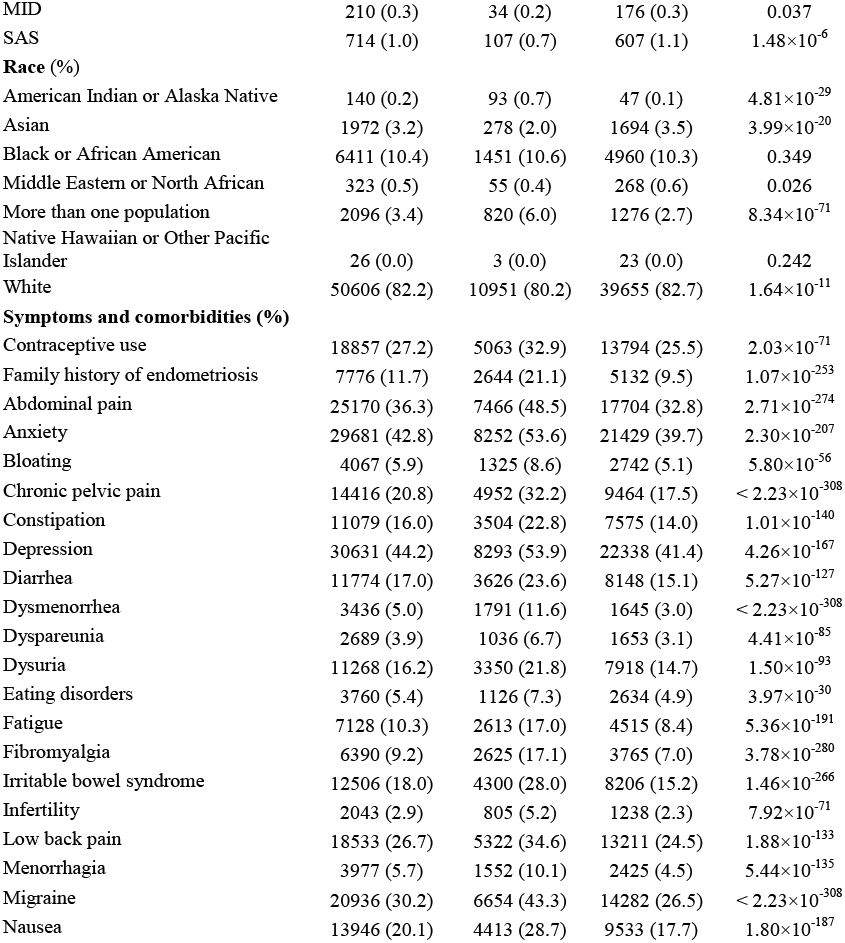
Characteristics of endometriosis cases and controls included in the current study.

|  | **Cases +**<br>**controls**<br>**(N=69,376)** | **Cases**<br>**(N=15,382)** | **Controls**<br>**(N=53,994)** | **p-value** |
| --- | --- | --- | --- | --- |
| **Age** (mean±SD) | 56.94 (16.25) | 57.53 (14.44) | 56.78 (16.72) | $3.42 \times 10^{-8}$ |
| **Body Mass Index** (mean±SD) | 29.31 (7.65) | 30.45 (7.79) | 28.98 (7.58) | $8.08 \times 10^{-93}$ |
| **Deprivation index** (mean±SD) | 0.31 (0.06) | 0.31 (0.06) | 0.31 (0.06) | 0.016 |
| **Standardized PRS** (mean±SD) | 0.02 (1.01) | 0.25 (1.01) | -0.05 (1.00) | $1.61 \times 10^{-237}$ |
| **Ancestry (%)** |  |  |  |  |
| AFR | 7322 (10.6) | 1773 (11.5) | 5549 (10.3) | $1.06 \times 10^{-5}$ |
| AMR | 7684 (11.1) | 1720 (11.2) | 5964 (11.0) | 0.631 |
| EAS | 1580 (2.3) | 216 (1.4) | 1364 (2.5) | $5.12 \times 10^{-18}$ |
| EUR | 51866 (74.8) | 11532 (75.0) | 40334 (74.7) | 0.501 |
| MID | 210 (0.3) | 34 (0.2) | 176 (0.3) | 0.037 |
| SAS | 714 (1.0) | 107 (0.7) | 607 (1.1) | $1.48 \times 10^{-6}$ |
| **Race (%)** |  |  |  |  |
| American Indian or Alaska Native | 140 (0.2) | 93 (0.7) | 47 (0.1) | $4.81 \times 10^{-29}$ |
| Asian | 1972 (3.2) | 278 (2.0) | 1694 (3.5) | $3.99 \times 10^{-20}$ |
| Black or African American | 6411 (10.4) | 1451 (10.6) | 4960 (10.3) | 0.349 |
| Middle Eastern or North African | 323 (0.5) | 55 (0.4) | 268 (0.6) | 0.026 |
| More than one population | 2096 (3.4) | 820 (6.0) | 1276 (2.7) | $8.34 \times 10^{-71}$ |
| Native Hawaiian or Other Pacific Islander | 26 (0.0) | 3 (0.0) | 23 (0.0) | 0.242 |
| White | 50606 (82.2) | 10951 (80.2) | 39655 (82.7) | $1.64 \times 10^{-11}$ |
| **Symptoms and comorbidities (%)** |  |  |  |  |
| Contraceptive use | 18857 (27.2) | 5063 (32.9) | 13794 (25.5) | $2.03 \times 10^{-71}$ |
| Family history of endometriosis | 7776 (11.7) | 2644 (21.1) | 5132 (9.5) | $1.07 \times 10^{-253}$ |
| Abdominal pain | 25170 (36.3) | 7466 (48.5) | 17704 (32.8) | $2.71 \times 10^{-274}$ |
| Anxiety | 29681 (42.8) | 8252 (53.6) | 21429 (39.7) | $2.30 \times 10^{-207}$ |
| Bloating | 4067 (5.9) | 1325 (8.6) | 2742 (5.1) | $5.80 \times 10^{-56}$ |
| Chronic pelvic pain | 14416 (20.8) | 4952 (32.2) | 9464 (17.5) | $< 2.23 \times 10^{-308}$ |
| Constipation | 11079 (16.0) | 3504 (22.8) | 7575 (14.0) | $1.01 \times 10^{-140}$ |
| Depression | 30631 (44.2) | 8293 (53.9) | 22338 (41.4) | $4.26 \times 10^{-167}$ |
| Diarrhea | 11774 (17.0) | 3626 (23.6) | 8148 (15.1) | $5.27 \times 10^{-127}$ |
| Dysmenorrhea | 3436 (5.0) | 1791 (11.6) | 1645 (3.0) | $< 2.23 \times 10^{-308}$ |
| Dyspareunia | 2689 (3.9) | 1036 (6.7) | 1653 (3.1) | $4.41 \times 10^{-85}$ |
| Dysuria | 11268 (16.2) | 3350 (21.8) | 7918 (14.7) | $1.50 \times 10^{-93}$ |
| Eating disorders | 3760 (5.4) | 1126 (7.3) | 2634 (4.9) | $3.97 \times 10^{-30}$ |
| Fatigue | 7128 (10.3) | 2613 (17.0) | 4515 (8.4) | $5.36 \times 10^{-191}$ |
| Fibromyalgia | 6390 (9.2) | 2625 (17.1) | 3765 (7.0) | $3.78 \times 10^{-280}$ |
| Irritable bowel syndrome | 12506 (18.0) | 4300 (28.0) | 8206 (15.2) | $1.46 \times 10^{-266}$ |
| Infertility | 2043 (2.9) | 805 (5.2) | 1238 (2.3) | $7.92 \times 10^{-71}$ |
| Low back pain | 18533 (26.7) | 5322 (34.6) | 13211 (24.5) | $1.88 \times 10^{-133}$ |
| Menorrhagia | 3977 (5.7) | 1552 (10.1) | 2425 (4.5) | $5.44 \times 10^{-135}$ |
| Migraine | 20936 (30.2) | 6654 (43.3) | 14282 (26.5) | $< 2.23 \times 10^{-308}$ |
| Nausea | 13946 (20.1) | 4413 (28.7) | 9533 (17.7) | $1.80 \times 10^{-187}$ |

### Prediction models

Race was the only variable which was excluded from the modeling due to high missingness (11.3%). All the other variables with missingness, BMI (1.8%), family history of endometriosis (4.1%), deprivation index (1.3%) could be imputed with MICE. Across all five prediction models, XGBoost achieved the highest discrimination, outperforming logistic regression and random forest (Figure 1, Supplemental Tables 2-6). The results below refer to cross-validated XGBoost performance, and models were compared using out-of-fold predictions on the same individuals. Discrimination increased as non-genetic information was added to the genetic and demographic base. The base model (Model 1: PRS, age, ancestry, and principal components) yielded an AUROC of 0.63 (Figure 1, Supplemental Table 2). Adding environmental and reproductive factors (Model 2) improved discrimination to 0.67 (vs Model 1, Δ=0.035, Holm-adjusted p=2.8×10^-23^; Figure 1, Supplemental Table 3), while adding symptom and comorbidity indicators produced a larger gain (Model 3, AUROC 0.71; vs Model 1, Δ=0.082, p = 2.8×10^-127^; Figure 1, Supplemental Table 4). The full model combining genetic, environmental, and symptom variables (Model 4) achieved the highest discrimination (AUROC 0.72; Figure 1, Supplemental Table 5), significantly exceeding both the symptom model (vs Model 3, Δ=0.012, p=7.6×10^-4^) and the environmental model (vs Model 2, Δ=0.059, p=5.9×10^-68^). Symptom information contributed the largest single increment: the symptom model alone (Model 3) outperformed the environmental model (Model 2) by a wide margin (Δ=0.047, p=1.9×10^-43^). Adding PRS × environment interaction terms (Model 5, AUROC 0.66; Figure 1, Supplemental Table 6) did not improve on the environmental model without interactions (Model 2, 0.67; Δ=-0.001, p=0.80), which was the only one of the ten pairwise comparisons that was not statistically significant after Holm correction, indicating that modeling interactions between PRS and environmental factors provided no additional predictive value (Supplemental Table 7). Overall discrimination was modest across all models (AUROC 0.63-0.72), and class weighting did not materially change discrimination relative to unweighted models (e.g., XGBoost AUROC 0.63 in both versions of Model 1). At the default probability threshold of 0.5, sensitivity was near zero because of the strong class imbalance, with models classifying almost all women as controls. At the Youden-optimal threshold, the best model (Model 4) reached a sensitivity of 0.67 and specificity of 0.65, with positive predictive value remaining low (~0.35), reflecting the low case prevalence (Figure 1, Supplemental Table 5).

**Figure 1.**
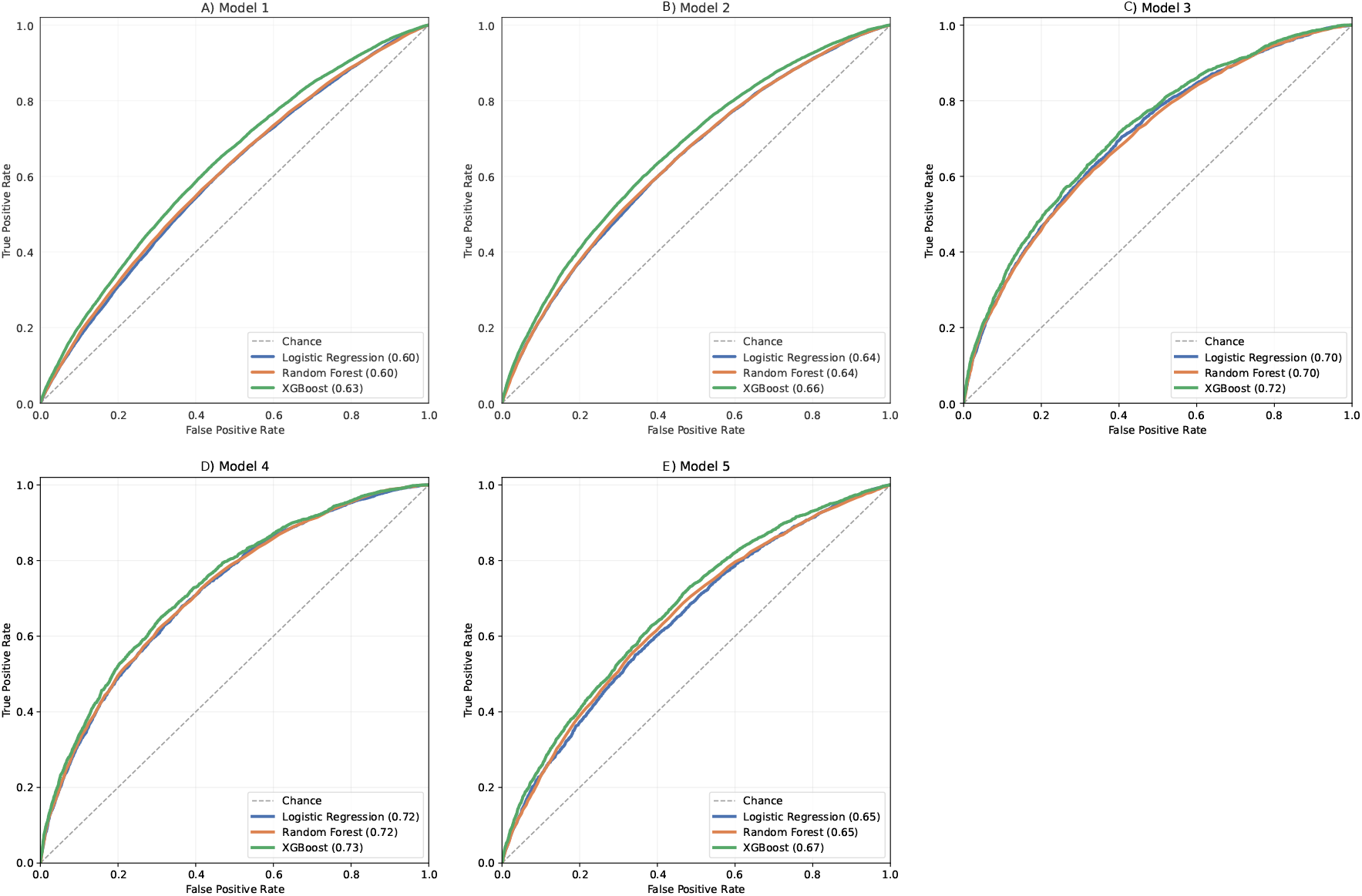
Receiver operating characteristic (ROC) curves for logistic regression, random forest, and XGBoost classifiers across the five nested prediction models (held-out test set). A) Model 1: PRS, age, genetic ancestry, and ancestry-specific principal components. B) Model 2: Model 1 plus environmental and reproductive factors (BMI, area deprivation index, contraceptive use, family history of endometriosis). C) Model 3: Model 1 plus symptom and comorbidity indicators. D) Model 4: full model combining genetic, demographic, environmental/ reproductive, and symptom/comorbidity features. E) Model 5: Model 4 plus PRS × environmental factor interaction terms. AUROC values for each classifier are shown in the legend of each panel. The dashed diagonal line indicates chance-level discrimination (AUROC = 0.50).

Permutation importance for the best-performing model showed that PRS and age were consistently the two largest contributors across models (Figure 2, Supplemental Tables 2-6). In the base model, PRS (mean decrease in AUROC 0.059) and age (0.057) accounted for essentially all discrimination, whereas ancestry and principal components contributed little. With environmental factors added (Model 2), family history of endometriosis (0.031), contraceptive use (0.019), and BMI (0.014) were the leading non-genetic predictors. When symptoms were included (Models 3 and 4), the strongest symptom contributors were migraine, irritable bowel syndrome, fibromyalgia, chronic pelvic pain, and dysmenorrhea, with age, PRS, and family history remaining among the top predictors overall. In the interaction model (Model 5), the PRS × environment terms each contributed little (mean decrease in AUROC ≤0.006), consistent with the absence of improvement over Model 2.

**Figure 2.**
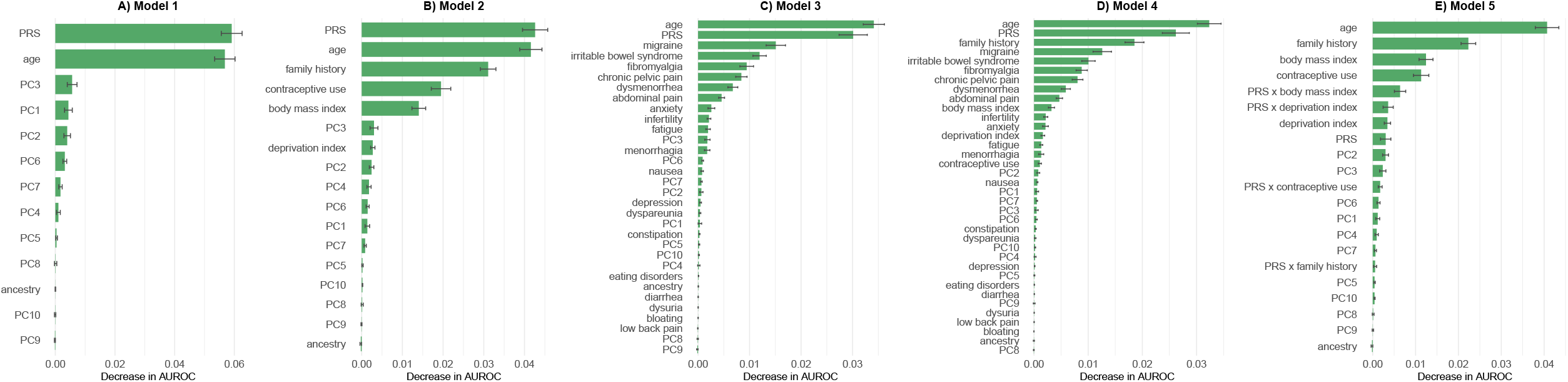
Permutation feature importance for the XGBoost classifier, the best-performing model in each comparison, across the five nested prediction models. A) Model 1 (PRS, age, genetic ancestry, and ancestry-specific principal components). B) Model 2 (Model 1 + environmental and reproductive factors (BMI, area deprivation index, contraceptive use, family history of endometriosis). C) Model 3 (Model 1 + symptom and comorbidity indicators). D) Model 4 (full model combining genetic, demographic, environmental/ reproductive, and symptom/comorbidity features). E) Model 5 (Model 4 + PRS × environmental factor interaction terms). Importance is defined as the mean decrease in AUROC across 20 random permutations of each feature’s values, with features ranked in descending order of importance within each panel.

## Discussion

In this study, we developed and compared hybrid risk-prediction models for endometriosis in the multi-ancestry *All of Us* cohort, integrating a genome-wide PRS with environmental, reproductive, and symptom information. Discrimination improved as clinical information was layered onto the genetic and demographic base, from an AUROC of 0.63 for PRS, age, ancestry, and principal components alone to 0.72 for the full model, with symptom and comorbidity data contributing the largest single increment. The PRS was among the strongest individual predictors, yet genetic and demographic information alone gave only modest discrimination, and modeling PRS × environment interactions provided no additional gain. Across all models, XGBoost performed best, though only marginally better than logistic regression and random forest, which is partially supported by previous observations: a previous study found no systematic performance benefit of machine learning over logistic regression across clinical prediction models^20^. However, machine learning methods frequently provide superior feature importance selection and ranking, capturing key predictors missed by logistic regression, handling multicollinearity more effectively, and offering superior ability to model complex, non-linear relationships^21,22^.

Our results extend and contextualize prior machine-learning models for endometriosis. Symptom-only questionnaire models have reported very high accuracy, up to an AUROC of 0.94^9^, but were developed in small, balanced samples recruited from disease-specific online communities (Facebook groups), evaluated only by internal cross-validation, and trained against undiagnosed rather than clinically confirmed controls, which inflates apparent performance and limits generalizability. In large, population-based cohorts more comparable to ours, discrimination has been more modest. A comorbidity-based model in approximately 628,000 Spanish primary-care records achieved an AUROC of 0.73^10^, essentially identical to our best model, but with a positive predictive value of only 1.5%, meaning that almost all positive predictions were false. A UK Biobank model combining lifestyle, physical, and diagnostic features reached an AUROC of 0.80^13^, but was reported using discrimination alone, without sensitivity, specificity, or predictive values, and adding candidate genetic variants did not improve prediction. Our study contributes a transparent and thorough evaluation: we report the full set of operating characteristics (sensitivity, specificity, positive and negative predictive value, balanced accuracy, F1 score, and confusion-matrix counts). These parameters, derived using a well-powered multi-ancestry PRS, apply to both default and Youden-optimized thresholds, and for both held-out and cross-validated predictions in an ancestrally diverse population.

A recurring challenge in endometriosis prediction is class imbalance. At the default probability threshold, our models, like others before, classified almost all women as controls, yielding high specificity and negative predictive value but negligible sensitivity. Such a model identifies who is unlikely to have endometriosis but does not flag cases. By evaluating class-weighted models and selecting an operating threshold that maximizes the Youden index, we obtained balanced specificity and sensitivity (Random forest, 0.67 and 0.65 for Model 4, respectively), making the models useful for case identification rather than only for ruling disease out. Positive predictive value nonetheless remained moderate rather than high, which is a consequence of operating at a single sensitivity/specificity trade-off point rather than of low disease prevalence, since endometriosis was in fact common in our sample (~22%). As previous authors have concluded, such models are best positioned as triage or prioritization tools that complement, rather than replace, clinical assessment.

The endometriosis PRS was one of the most influential predictors across models, ranking at or near the top of the permutation-importance analyses alongside age, and remaining a leading feature even after symptom and comorbidity variables were added. This contrasts with prior work in which small sets of candidate GWAS variants contributed negligibly, and indicates that a well-powered, genome-wide multi-ancestry PRS carries meaningful individual-level signal. At the same time, genetic and demographic information alone provided only modest discrimination (AUROC 0.63), and the largest incremental gains came from symptom and comorbidity data (0.72). PRS is therefore an important but not sufficient component of endometriosis risk prediction, most valuable when combined with clinical information rather than used in isolation. Consistent with this, modeling interactions between PRS and environmental factors did not improve discrimination, suggesting either insufficient power to detect it or indeed true absence of meaningful interaction, as several important factors such as endocrine disruptors^23^ are not available in biobanks.

Several paths could improve these results. First, incorporating additional data modalities, such as circulating inflammatory and hormonal proteins/biomarkers, miRNA, metabolomic profiles, imaging features, medication history, and longitudinal data from menstrual-tracking applications/wearables may capture disease signal not reflected in coded diagnoses and symptoms. Second, moving beyond a binary case/control outcome to predict clinically meaningful subtypes (e.g., ovarian, deep infiltrating, or peritoneal disease, and co-occurring adenomyosis)^24^, disease severity^12,24^, or the symptom-based phenotypes we previously identified^3^ could yield more actionable models, since risk factors and biology may differ across these presentations. Third, temporal modeling that uses only information available before diagnosis and that distinguishes antecedent risk factors from downstream consequences of the disease would better serve the goal of shortening diagnostic delay. Finally, external validation in independent, diverse cohorts, together with assessment of calibration and clinical net benefit (e.g., decision-curve analysis), will be essential before any such model could inform practice.

This study has limitations. Case ascertainment relied on EHR codes and self-report, which are subject to misclassification and underdiagnosis, and the analysis was cross-sectional. Because symptom and comorbidity codes come from the same EHR system as the outcome, people who interact with healthcare more often will accumulate more of both, independent of true biological association. In this case the models are better understood as diagnostic-associative than as prospective screening tools. The PRS was derived predominantly from European-ancestry summary statistics, likely limiting its transferability to other groups. Socioeconomic status was captured only through a zip-code-level deprivation index, which may not fully reflect individual-level socioeconomic circumstances. Environmental exposures were also incompletely captured: variables such as endocrine-disrupting chemical exposure and detailed dietary patterns, which have been implicated in endometriosis risk, were not available in this dataset and would be valuable additions in future work. Finally, although *All of Us* is large and ancestrally diverse, it is a US-based, volunteer-based, opt-in cohort and not a random population sample. External replication is needed to establish generalizability.

In summary, the PRS ranked among the most influential individual predictors, with symptoms and comorbidities providing the largest incremental discrimination. Integrating polygenic, environmental, reproductive, and symptom information yielded only modest discrimination for endometriosis, with no benefit from PRS-environment interactions. By reporting complete operating characteristics across a diverse population within a common framework, our work provides a realistic benchmark for hybrid endometriosis prediction and highlights the data types and study designs most likely to advance it.

## Data availability statement

This study used individual-level data from the AoU Research Program, which are not publicly downloadable. De-identified electronic health record and survey data can be accessed by approved researchers through the AoU Researcher Workbench (https://workbench.researchallofus.org) after institutional registration, completion of the required training, and agreement to the Data User Code of Conduct. All derived summary data resulting from the analyses are provided in the Supplementary material accompanying this manuscript.

## Acknowledgements

We gratefully acknowledge All of Us participants for their contributions, without whom this research would not have been possible. We also thank the National Institutes of Health’s All of Us Research Program for making available the participant data examined in this study. The AoU Research Program is supported by the National Institutes of Health, Office of the Director: Regional Medical Centers: 1 OT2 OD026549; 1 OT2 OD026554; 1 OT2 OD026557; 1 OT2 OD026556; 1 OT2 OD026550; 1 OT2 OD 026552; 1 OT2 OD026553; 1 OT2 OD026548; 1 OT2 OD026551; 1 OT2 OD026555; IAA: AOD 16037; Federally Qualified Health Centers: HHSN 263201600085U; Data and Research Center: 5 U2C OD023196; Biobank: 1 U24 OD023121; The Participant Center: U24 OD023176; Participant Technology Systems Center: 1 U24 OD023163; Communications and Engagement: 3 OT2 OD023205; 3 OT2 OD023206; and Community Partners: 1 OT2 OD025277; 3 OT2 OD025315; 1 OT2 OD025337; 1 OT2 OD025276.

## Author contributions

D.K. designed the study. O.G. and D.K. conducted statistical analyses. O.G. and D.K. interpreted the results. D.K. wrote the original draft of the manuscript. All authors provided comments and revised the manuscript. D.K. provided funding and supervised the analyses.

